# “Informative Use of Cycle-Threshold Values to Account for Sampling Variability in Pathogen Detection”

**DOI:** 10.1101/2023.03.21.23287544

**Authors:** Aidan M. Nikiforuk, Agatha N. Jassem

## Abstract

Nucleic acid amplification tests, like real-time polymerase chain reaction, are widely used for pathogen detection; however, their interpretation rarely accounts for sampling variability. Instead, cycle threshold values are often categorized reducing precision. We describe how pathogen cycle threshold values can be normalized to endogenous host gene expression to correct for sampling variability and compare the validity of this approach to standardization with a standard curve. Normalization serves as a valid alternative to standardization, does not require making a standard curve, increases precision, accounts for sampling variability, and can be easily applied to large clinical or surveillance datasets for informative interpretation.

## Introduction

Nucleic acid amplification tests like reverse transcription quantitative real-time polymerase chain reaction (RT-qPCR) are routinely used in clinical laboratories for pathogen detection [1]. The prototypical RT-qPCR assay includes two primer and probe sets (targets) for a specific pathogen and one primer probe set (target) for an endogenous host gene. Results are interpreted from a range of cycle-threshold (Ct) values, samples with a Ct value below a certain predetermined cut-off are categorized as positive [2]. Categorizing Ct values transforms a quantitative measurement into a categorical one limiting precision of the measurement which may lead to misinterpretation. Yet, Ct values are broadly interpreted categorically because, it provides a simpler alternative to using them as numerical value, which requires making a standard curve and additional analysis.

To encourage more informative use of Ct values as a numerical value, we investigate using relative gene expression (normalization) in lieu of a standard curve (standardization) for interpretation [3]. Normalization has the benefits of re-purposing the host gene target, accounting for sampling variability by measuring the ratio of the pathogen target(s) to host target, it can be performed retrospectively and automated for large clinical or surveillance datasets.

## Methods

To validate relative gene expression as a quantitative way to interpret Ct values we used a dataset of n = 212 clinical test results from persons who tested positive for COVID-19 from 24/3/2020 to 9/5/2020. Specimens from persons seeking a diagnostic test for SARS-CoV-2 infection were collected by nasopharyngeal swab and nucleic acid extraction was performed using the Viral RNA isolation kit on the MagMAX-96™ platform (ThermoFisher). Host (*RNaseP*) and viral gene (*Envelope, Nucleocapsid*) targets were assayed by RT-qPCR, as previously described. A 9-replicate, 5-fold, 1:10 dilution of SARS-CoV-2 synthetic RNA was used to make a standard curve and Ct values were transformed to log10 GE/mL using simple linear regression[4]. Relative gene expression was calculated between the viral Envelope (*E*) and *RNaseP* targets using a derivation of the method proposed by Livak and Schmittgen (2^−ΔCt^) [5]. Pearson’s correlation and simple linear regression were used to estimate the relationship between crude, normalised and standardized RT-qPCR measurements. Data analysis was performed using R Statistical Software Version 4.1.0, p-values of less than α = 0.05 were considered statistically significant.

## Results

To illustrate how normalizing the pathogen gene target(s) to host gene target accounts for sampling variability we made a scatterplot of normalized SARS-CoV-2 viral load to crude viral *E* gene Ct (Figure 1). At a single Ct value (e.g. Ct 15) the quantifiable amount of SARS-CoV-2 genome differs by 5-fold, which corresponds to 10,000 viral genomes or a 4-log change. This difference was undetectable without correcting for sampling variability and serves as an example of how interpretation of untransformed Ct values may lead to information bias and incorrect inference.

**Figure 1:**
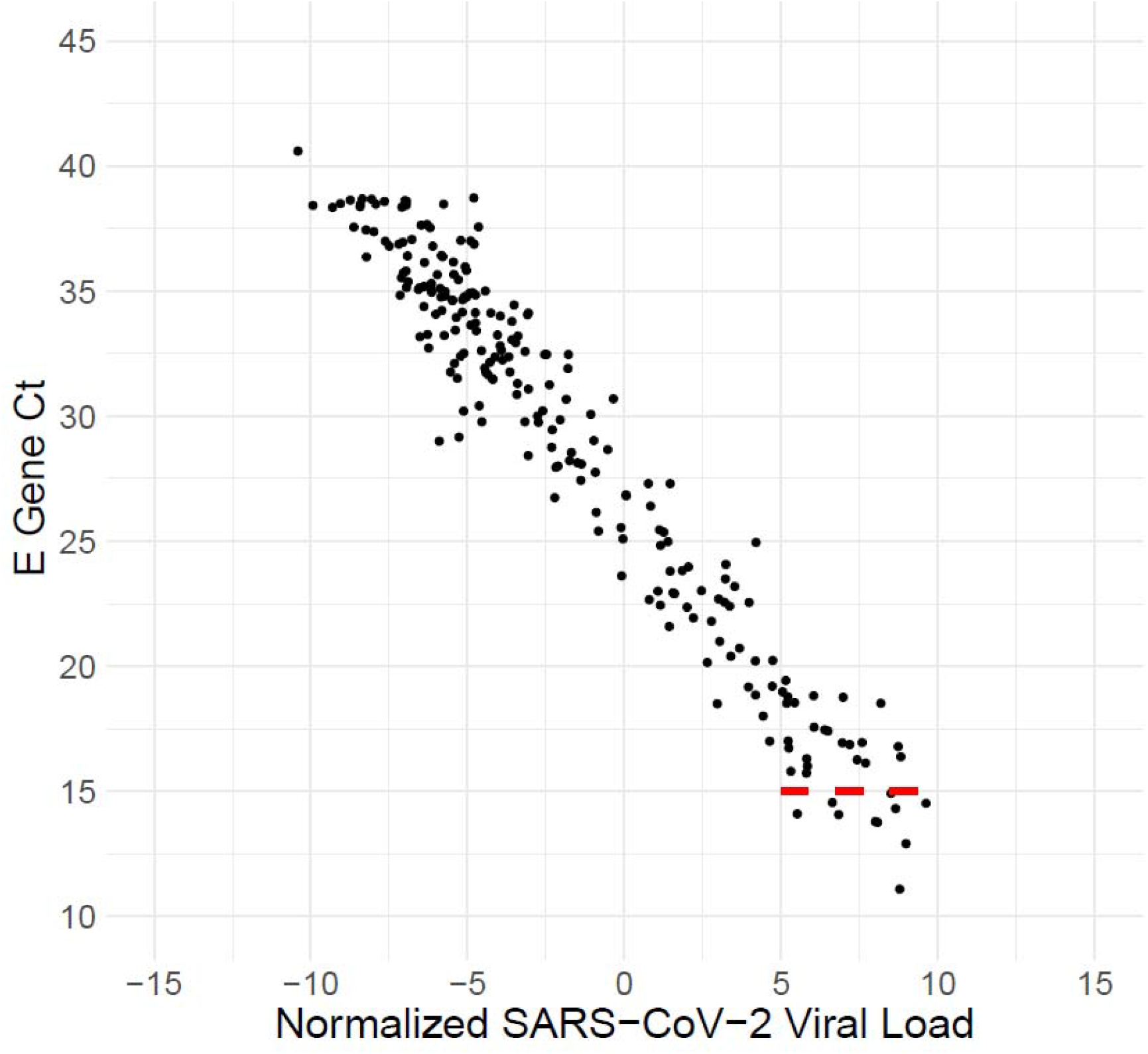
Scatterplot Visualizing the Linear Relationship Between Normalized SARS-CoV-2 Viral Load and SARS-CoV-2 E gene Cycle-Threshold Value, for n = 212 Nasopharyngeal Specimens (PCC, r = -0.970, P < 0.001). Scatterplot of normalized SARS-CoV-2 viral load and *Envelope* gene (*E*) cycle threshold values (Ct) assayed by RT-qPCR in n = 212 nasopharyngeal specimens collected from people tested for COVID-19 in British Columbia. The transformed and untransformed variables show a strong negative relationship, quantified by Pearson’s correlation coefficient (r = -0.970, P < 0.001). The red segmented line shows that at a constant E gene Ct value of 15, SARS-CoV-2 viral load may vary by a 5-fold difference which corresponds to 10,000 viral genomes. This dramatic difference would not have been detectable without adjusting for sampling variability in the normalization process; therefore, Ct values alone possess little quantitative information regarding viral load.

We then compared normalized SARS-CoV-2 viral load to standardized viral load to test if relative gene expression serves as a valid alternative to making a standard curve (Figure 2). Normalized viral load has a strong positive linear relationship with standardized measurements and variability seems homogenous across the range of observed values, indicating comparability (SLR, R^2^ = 0.942, P < 0.001).

**Figure 2:**
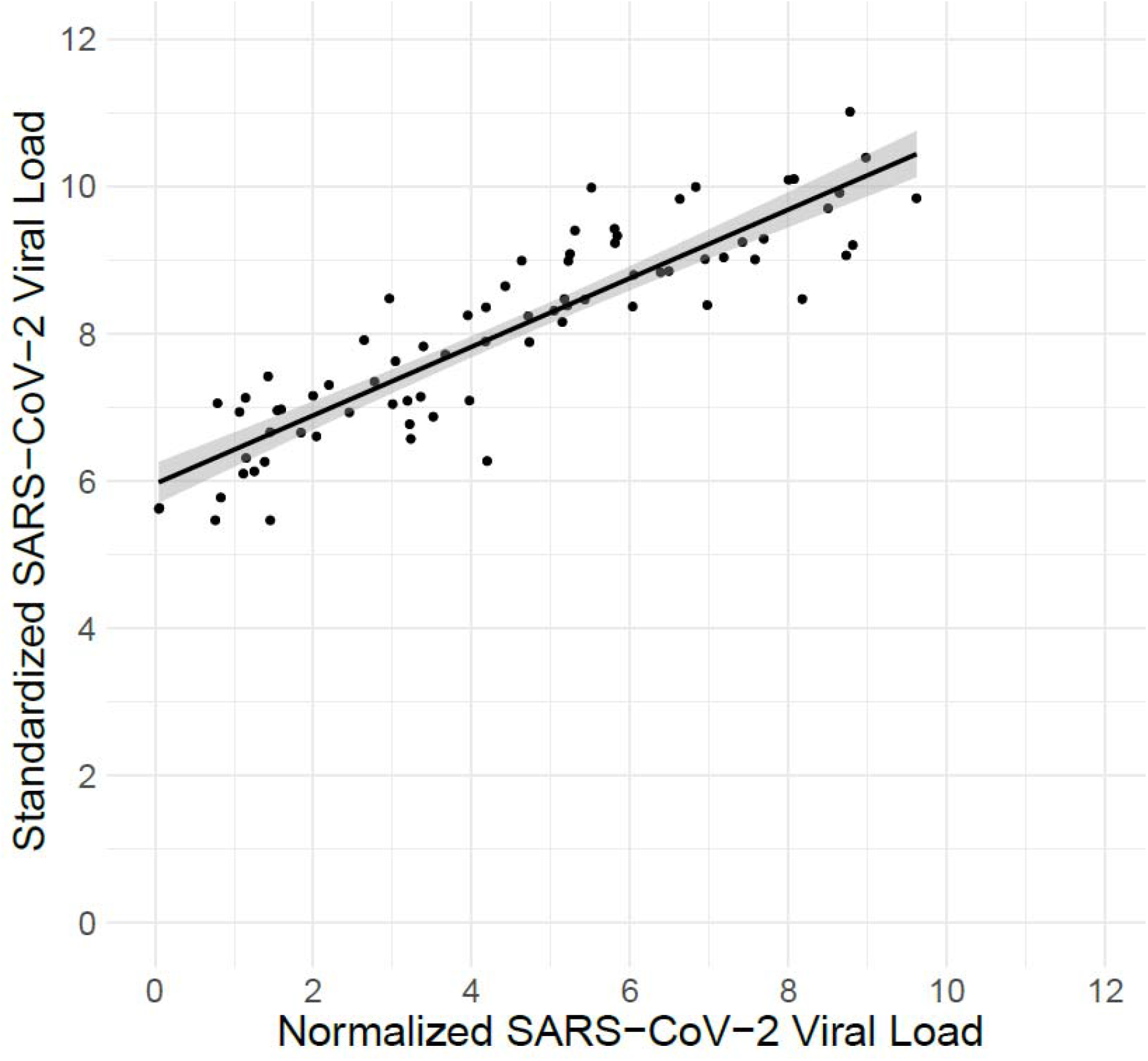
Linear Relationship between normalized SARS-CoV-2 viral load and SARS-CoV-2 E gene Cycle-Threshold Value, for n = 212 Nasopharyngeal Specimens (SLR, R^2^ = 0.942, P < 0.001). Scatterplot of normalized and standardized SARS-CoV-2 viral load assayed by RT-qPCR in n = 212 nasopharyngeal specimens collected from people tested for COVID-19 in British Columbia. The two variables show a strong positive relationship and shared variation, quantified by simple linear regression (R^2^ = 0.942, P < 0.001). Black line and shading show the model fit and 95% confidence interval.

## Discussion

We demonstrate that using relative gene expression to normalize pathogen load quantifies specimen sampling variability and serves as a valid alternative to analysis using a standard curve. Normalization has the benefits of: repurposing host gene targets, accounting for sampling variability (reducing information bias), more informative use of Ct values as a numeric measure than a categorical one and can be retrospectively performed on large clinical datasets. Relative gene expression makes the assumption that the target host gene selected as an endogenous control possess equivalent inter-individual expression, which may not always be the case in those infected with a pathogen.

Given the well document disadvantages of interpreting Ct values categorically or as a crude numeric measure for pathogen detection[6], we recommend that clinicians consider making more informed use and account for sampling variability by normalization as described.

## Data Availability

The de-identified participant data that support the findings of this study are available from the British Columbia Ministry of Health. Restrictions apply to the availability of these data, which were used under agreement at the British Columbia Center for Disease Control for this study.

## Abbreviations

RT-qPCR: real-time quantitative polymerase chain reaction
Ct: cycle threshold
RNaseP: Ribonuclease P
E: Envelope
PCC: Pearson’s correlation coefficient
SLR.: simple linear regression

## Ethical Approval

Ethical approval for the study was obtained from the University of British Columbia human ethics board (H20-01110). Written informed consent was not required as per ethics board approval. Participant data was de-identified prior to analysis; the results of this non-interventional observational study were not linked back to any identifying patient records. The study was deemed to be of minimal risk.

## Author Contributions

A.M. Nikiforuk: Conceived, designed, and performed the analysis, wrote the manuscript, reviewed, and edited the manuscript. A.N. Jassem: Conceived the analysis, discussed the results, reviewed, and edited the manuscript.

